# ERBB2 R599C variant is associated with left ventricular outflow tract obstruction defects in human

**DOI:** 10.1101/2023.11.17.23297969

**Authors:** M Ampuja, S Selenius, I Paatero, I Chowdhury, J Villman, M Broberg, A Ranta, T Ojala, JX Chong, M Bamshad, JR Priest, M Varjosalo, R Kivelä, E Helle

**Author notes:** These authors have contributed equally (R Kivelä, E Helle).

## Abstract

**Background and aims:** Non-syndromic congenital heart defects (CHD) are occasionally familial and left ventricular out flow tract obstruction (LVOTO) defects are among the subtypes with the highest hereditability. The aim of this study was to evaluate the pathogenicity of a heterozygous *ERBB2* variant R599C identified in three families with LVOTO defects.

**Methods:** Variant detection was done with exome sequencing. Western blotting, digital PCR, mass spectrometry (MS), MS-microscopy and flow cytometry were used to study the function of the *ERBB2* variant R599C. Cardiac structure and function were studied in zebrafish embryos expressing human *ERBB2* WT or R599C. Patient-derived human induced pluripotent stem cell cardiomyocytes (hiPS-CM) and endothelial cells (hiPS-ECs) were used for transcriptomic analyses.

**Results:** While phosphorylation of the ERBB2 R599C receptor was not altered, the variant affected dramatically the binding partners of the protein and lead to mislocalization of ERBB2 from plasma membrane to ER and mitochondria. Expression of human ERBB2 R599C in zebrafish embryos resulted in cardiomyocyte hypertrophy, increased cardiac wall thickness, and impaired fractional shortening, demonstrating that the mutant receptor induces functional and structural defects during heart development. Transcriptomic analyses of hiPS-ECs and hiPS-CMs from a patient with the R599C variant indicated aberrant expression of genes related to cardiovascular system development and abnormal response to oxidative stress in both cell types.

**Conclusion:** The heterozygous variant *ERBB2* R599C leads to abnormal cellular localization of the ERBB2 receptor inducing structural changes and dysfunction in the zebrafish embryo heart. This evidence suggests ERBB2 as a novel disease gene for CHD.

## Introduction

Left ventricular outflow tract obstruction (LVOTO) is a subgroup of congenital heart defects (CHD) affecting the left side of the heart – the mitral valve, the left ventricle, the aortic valve and the aorta. The severity of LVOTO defects range from the often initially asymptomatic bicuspid aortic valve (BAV), to complex defects, such as hypoplastic left heart syndrome (HLHS), representing one of the most severe forms of CHD. In HLHS, mitral and aortic stenosis or atresia combined with left ventricular hypoplasia result in the left side of the heart incapable of adequately supporting the systemic circulation. This necessitates palliative procedures, ultimately leading to the establishment of single ventricle physiology.

Non-syndromic CHD are often hereditary, and LVOTO defects are among the subtypes with the highest hereditability - they have been shown to be associated with a 20% incidence of CHD in the first-degree relatives (1, 2). Around 5-15% non-syndromic CHD are estimated to be monogenic, and it has been suggested that the majority of CHD are oligogenic with two or more predisposing genetic variants contributing (3, 4), or multifactorial, occurring due to a combination of genetic and environmental risks (5–8). Indeed, genetic susceptibility in the form of rare risk alleles with modest effect sizes have been observed in genome wide association (GWAS) studies for LVOTO (9, 10) as well as for other CHD (9, 11, 12). Out of environmental risks, maternal diabetes, obesity, advanced maternal age, maternal hypertension, maternal medications, and certain viral infections during early pregnancy are the most well documented (5, 13–18).

Identifying genes associated with CHD and LVOTO defects even in familial cases is complicated by their varied inheritance patterns, reduced penetrance and variable expressivity (19, 20). Monogenic forms with autosomal dominant inheritance due for example, to truncating variants in *NOTCH1,* have been identified as causal in some LVOTO families with multiple affected members (21–24).

Identifying new CHD genes will provide tools for prenatal and genetic counseling and provide cues to aid in further unraveling the complex molecular mechanisms of cardiac development and the cellular level events leading to CHD (9, 10). The relatively genetically homogenous Finnish population provides an excellent opportunity to identify disease-related variants compared to more heterogeneous populations (25) - especially as there are regional differences in the prevalence of certain CHD subtypes within Finland (26).

To gain further insight into the genetic etiology of LVOTO and human cardiac development, we conducted exome sequencing in Finnish LVOTO subjects and families with multiple affected members to identify new LVOTO associated loci. We identified an ultra-rare variant in the *ERBB2* gene that segregated with disease in three unrelated families. Functional analysis of the variant supports its causality for disease and highlights the role of ERBB2 in cardiac development, previously demonstrated only in animal models (27–31).

## Materials and Methods

### Exome sequencing, comparison to genome database

We recruited a cohort of 141 patients (81 singletons, 51 trios and 9 families with multiple affected members, total sequenced 275) with left ventricular outflow tract obstruction (LVOTO) defects from Helsinki University Children’s Hospital. Patients with a known or suspected syndrome or patients with extracardiac congenital malformations were not included. Exome sequencing was performed by the University of Washington Center for Mendelian Genomics Seattle, USA, and Blueprint Genetics Ltd. All persons sequenced were of self-reported Finnish ancestry.

### Variant calling

After filtering out duplicates and outliers based on genome quality, we used 275 hg19 aligned BAM files from both Blueprint Genetics (BPG, 173 exomes) and University of Washington (UW-CMG, 102 exomes) for variant calling using Freebayes v1.3.1 (32). We used the following settings for freebayes: --min-mapping-quality 20 --min-base-quality 20 --min-alternate-count 20 --min-alternate-fraction 0.2 -- no-partial-observations. The samples were compared against the gnomAD 2.1 database for detecting potential novel variants. We identified extremely rare MAF<0.0001 and novel variants present in a minimum of three probands in known CHD genes and genes known to be associated with cardiac development (33).

### Human induced pluripotent stem (hiPS) cell lines

Four human induced pluripotent stem cell lines (HEL47.2, HEL24.3, HEL46.11, HEL149.2) were obtained from the Biomedicum Stem Cell Center Core Facility. The cell lines were created by using retroviral/Sendai virus transduction of Oct3/4, Sox2, Klf4, and c-Myc, as described previously (34, 35). In addition, the hiPS line K1 was a kind gift from Prof. Anu Wartiovaara group. hiPSCs were maintained in Essential 8 media (A1517001, Thermo Fisher Scientific) on thin-coated Matrigel (354277, dilution 1:200; Corning). The cells were passaged using EDTA. The cells were routinely tested for mycoplasma with MycoAlert Mycoplasma Detection Kit (Lonza, LT07-218).

### hiPS-cardiomyocyte (CM) differentiation

Differentiation of hiPS cells to CMs was done as described previously by us (36). Briefly, consecutive treatment with Wnt pathway activator CHIR and inactivator IWR-1 was used to start differentiation. The cells were maintained in RPMI medium with B27 supplement, either with glucose, or without glucose and added lactate.

### hiPS-endothelial cell (EC) differentiation

Endothelial cell differentiation was performed as described previously by us (37). Briefly, BPEL media with BMP4, Activin A and CHIR was used to start differentiation. On day 3, media was changed to BPEL with VEGF and Activin A. ECs were sorted with CD31 beads between days 7-9 and immediately used for experiments.

### CM processing for scRNA-seq

Four LVOTO (HEL149.2, HEL169.4, HEL218.6, HEL216.6) and four healthy control (HEL47.2, HEL24.3, HEL46.11, K1) iPS cell lines were differentiated to CMs and at 38 or 39 days old (except HEL47.2 which was 27 days old) processed for scRNA-seq as described earlier (36, 37). Briefly, the cells were counted and washed with 0.04% BSA in PBS. Patient cells were combined with equal numbers of cells to one tube, and equal numbers of the four control cell lines were combined to one tube. The cells were taken to the Institute for Molecular Medicine Finland (FIMM) for processing with the 10X Genomics Single Cell Protocol. To identify the HEL149.2 sequencing data from the pooled sample, we utilized the freebayes v1.3.1 (38) software to call variants from the exomes from the four LVOTO patients. The called variants were used in the demuxlet software (39) to identify which cells belonged to the patient with R599C variant (HEL149.2) in a combined scRNA-seq dataset The cells from the other three LVOTO patients were not used for experiments. The identified cells/sample groups were further analysed, and the HEL149.2 data were compared to data from the pooled control data.

### scRNA-seq bioinformatics

Analysis was performed in R-studio using Seurat version 4.3.0. Cells with a minimum of 200 features, maximum of 8000 features and < 30% mitochondrial content were included. Normalization was carried out using ‘NormalizeData’ and ‘ScaleData’ and ‘FindVariableFeatures’. To integrate the data we used ‘SelectIntegrationFeatures’, FindIntegrationAnchors’ and ‘IntegrateData’ functions. Principal component analysis (PCA) was carried out on highly variable genes (dims=1:30, Resolution=0.2), these were used for uniform manifold approximation (UMAP). FindAllMarkers function was used to find differentially expressed genes. Established markers on The Human Protein Atlas and published literature were used to annotate cell types.

### EC processing for RNA-seq, RNA-seq

The patient line (HEL149.2) with the ERBB2 mutation and three control cell lines were used (HEL47.2, HEL24.3, K1). EC differentiation was done twice for each cell line, and one sample was collected from each differentiation when confluent (day 7-9). The ECs were sorted using magnetic beads with an antibody against CD31 (130-091-935, Miltenyi Biotec) according to the manufacturer’s protocol. The cells were then counted and collected in RA1 lysis buffer before extraction. For extraction, we used Nucleospin RNA Plus Extraction kit (740984, Macherey-Nagel). RNA-seq was performed at Biomedicum Functional Genomics Unit (FuGU) with Illumina NextSeq sequencer (Illumina, San Diego, CA, United States) using llumina Stranded Total RNA Prep with Ribo-Zero Plus. RNA-seq was performed as single-end sequencing for 75 bp read length.

### RNA-seq bioinformatics

The data was analysed using Chipster software (40). Differential expression was calculated using DEseq2. Genes with adjusted p-values of <0.05 for the log2fold-change were considered significant. Only genes with log2Fold-change on >0.25 were included. DAVID Bioinformatics Resources 6.8 was used for gene ontology (GO) analysis (41, 42).

### Flow cytometry

CMs were detached and counted. Of each sample, 1.1 million cells were aliquoted, centrifuged and suspended in 100 µl of cell staining buffer (2% FBS, 0.09% sodium azide in PBS). 5 µl of PE-conjugated ERBB2 antibody (BioLegend, 324406) was added to each sample. After 30 min of incubation at RT protected from light, the cells were centrifuged, washed with 1 ml of staining buffer and finally suspended in 500 µl of staining buffer. The cells were then analysed at the Flow Cytometry Core in Biomedicum with BD Influx cell sorter.

### Zebrafish transgenesis and videomicroscopy

Vectors containing ERBB2 and ERBB2 R599C as well as empty vectors with mCherry under myocardium-specific cmlc2-promoter were injected into 1-4 cell stage zebrafish casper or kdrl:EGFP embryos using Nanoject II microinjector (Drummond Scientific). One day after injection, the dead & malformed embryos were removed, and embryos cultured in E3-medium supplemented with 0.2mM PTU until analysed. Injected embryos displaying mCherry fluorescence were selected for phenotypic analyses. Embryos were anesthetized with MS-222 (200mg/L) and allowed to acclimatize to room temperature for at least 30 minutes before videos were taken at four dpf using Zeiss AxioZoom stereomicroscope and imaging at 30fps frame rate. Five to ten second movies were recorded from each embryo. Videos were analysed with FIJI software (43). Statistical analyses were done with GraphPad Prism 8 software.

### Responsible science

This study has been designed according to The Helsinki Declaration and the Conventions of the Council of Europe on human rights. The study protocols have been approved by the Ethical committee of Helsinki University Hospital District. Written informed consent has been obtained from all study participants over 6 years of age and from their parents/guardians if the participants are minors.

## Results

A missense SNV in *ERBB2* (chr17:37873630 C>T, Arg599Cys (hg19), rs369903296) was identified in three unrelated probands with LVOTO defects (Figure 1A, B). All three of these probands were familial cases with multiple affected members. We then identified the presence of the variant in the exome data of the family members of two probands where familial exomes were available, and did targeted Sanger sequencing of the family members of the third proband who was initially recruited as a singleton. In addition to the probands, the variant was found in all other affected family members, and altogether two unaffected family members (not shown to minimize identifiable data), suggesting reduced penetrance. One variant carrier in Family 3 has not been studied by echocardiogram.

**Figure 1:**
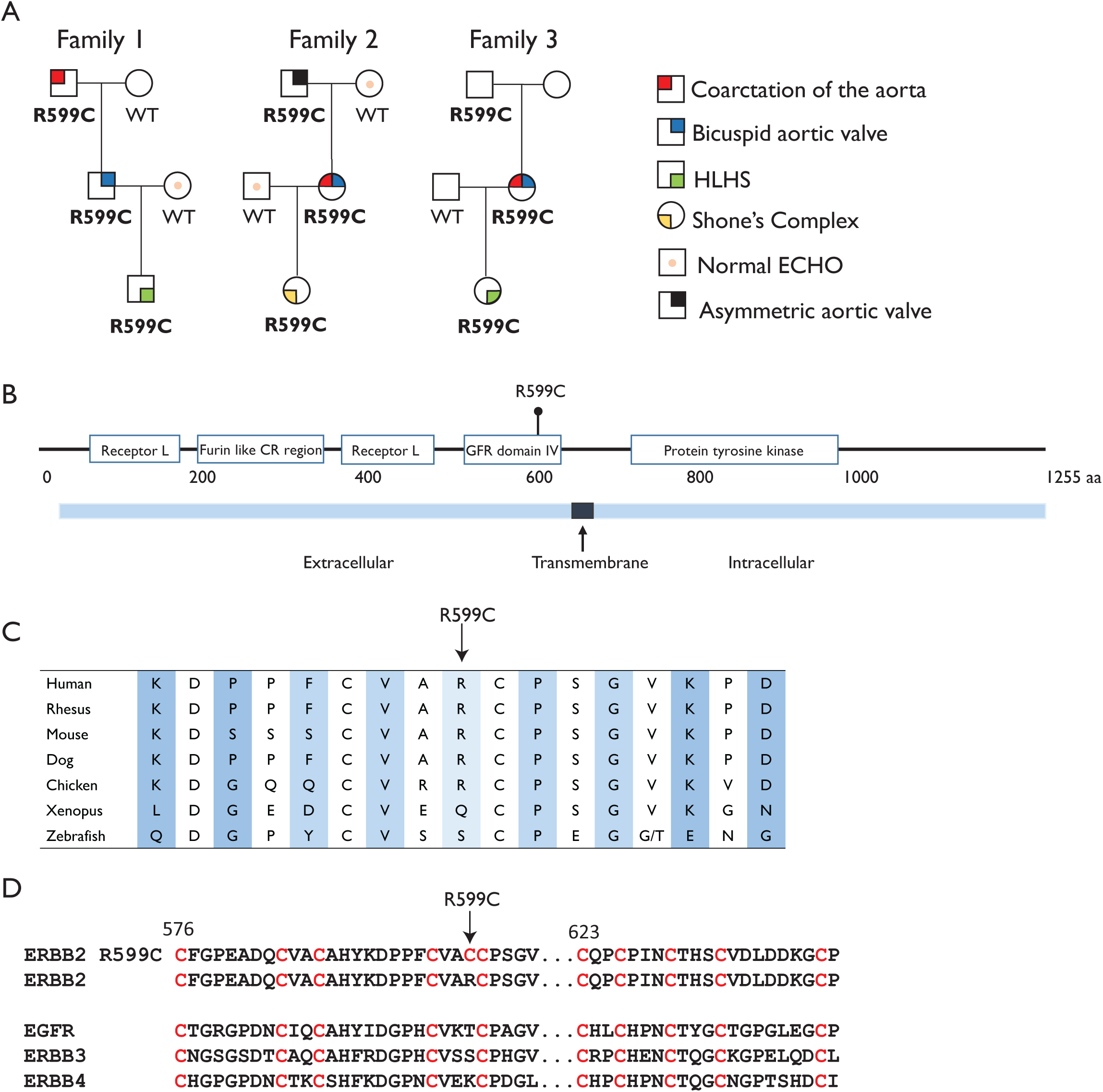
A) Pedigrees of the three affected families. Siblings are not included in the pedigree to minimize identifiable data. B) The R599C variant is located in the extracellular GFR domain IV of the ERBB2 protein. C) ERBB2 protein sequence conservation around the variant site. D) Protein sequence alignment of the four members of the EGFR family and the ERBB2 R599C mutant showing conservation of the cysteine sites in the four WT receptors, and addition of an extra cysteine in the ERBB2 R599C mutant.

The chr17:37873630 C>T variant is present in heterozygotic form in two subjects in GnomAD (v2.1.1, one Finnish European and one non-Finnish European), with a total allele frequency of 0.000007075 across all populations. The variant is not reported in Clinvar. It is predicted to be possibly damaging in Polyphen and deleterious in SIFT, and it has a CADD score of 29.9 (GRCh37 v1.6). Multiple sequence alignment shows high conservation of the variant locus *ERBB2* in multiple species (Figure 1C). In addition, the cysteine residue locations of the variant region are conserved in the four EGFR family members (Figure 1D).

### The ERBB2 R599C receptor is functional

As the variant results in an extra cysteine residue in the extracellular part of the ERBB2 receptor close to the transmembrane region, we hypothesized that this could induce changes in the receptor function. We first investigated the functionality of the mutant ERBB2 receptor by transfecting Cos7 cells with lentivirus containing either wild type (WT) *ERBB2* vector or *ERBB2* R599C vector. Western blot results show that ERBB2 R599C can be phosphorylated at Tyr1284 at least at the same level as the ERBB2 WT, or even more (Figure 2A, B).

**Figure 2:**
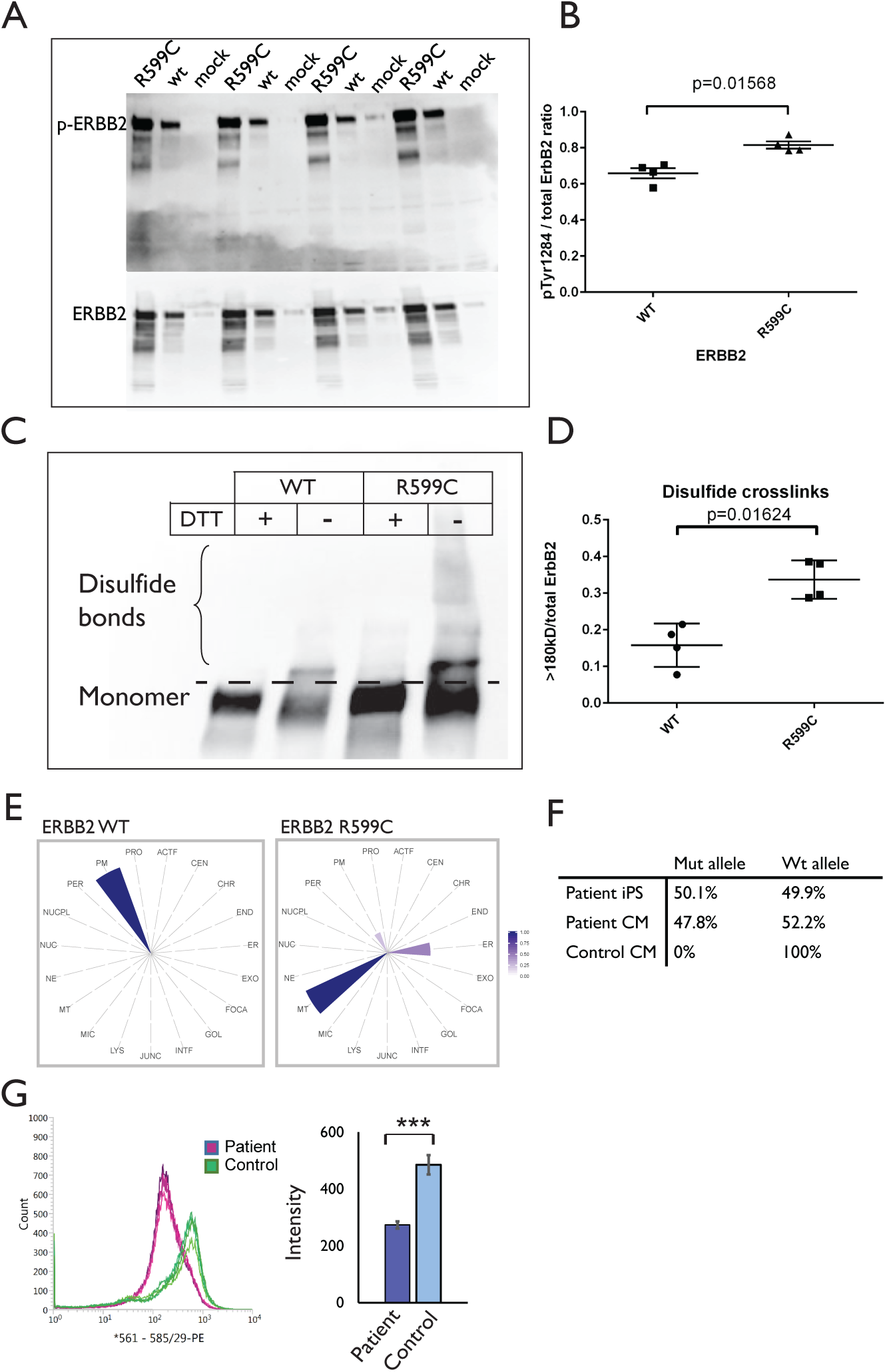
A) Cos-7 cells were transfected with either *ERBB2* WT plasmid, *ERBB2* R599C plasmid or empty plasmid (mock). Western blot with an antibody against p-ERBB2 and ERBB2 shows phosphorylation level of the receptor. B) Quantification of the phosphorylation levels show higher phosphorylation levels of the ERBB2 R599C receptor. C) Cos-7 cells were transfected with either *ERBB2* WT or *ERBB2* R599C plasmid. Leaving out the reducing agent (DTT) from the Western blot reveals the ERBB2-protein complexes that are larger due to intact disulfide bonds. D) Quantification of the disulfide bonds from the Western blot show higher amount of disulfide crosslinks in for the ERBB2 R599C receptor. E) MS Microscopy shows differences in the predicted cellular location for the ERBB2 WT and ERBB2 R599C receptor. F) Digital PCR confirms that the variant allele accounts for half of the *ERBB2* expression levels in patient derived hiPSCs and hiPS-CMs whereas the hiPS-CMs from a control subject (HEL47.2) has 100% expression of the WT allele. G) Flow cytometry with a PE-conjugated antibody that recognizes the extracellular part of ERBB2, and quantification of the intensities in the patient hiPS-CMs and two control hiPS-CMs (experiment repeated three times) indicate reduced levels of ERBB2 in hiPS-CMs from the patient with the *ERBB2* R599C variant. *** p < 0.005

### The ERBB2 R599C receptor has different interaction partners compared to ERBB2 WT

We next hypothesized that the change from arginine to cysteine, which introduces a free cysteine in the transmembrane domain of the receptor, may affect dimerization, binding, and/or localization of the receptor.

We first tested this by gel electrophoresis and Western blotting. Cos7 cells were transfected with *ERBB2* WT or *ERBB2* R599C plasmids and the cell lysates were run in gel electrophoresis with and without reducing agent DTT. Protein samples without DTT retain their disulfide bonds. The results show that there are significantly more ERBB2 disulfide bonds in the *ERBB2* R599C transfected cells compared to the *ERBB2* WT cells (Figure 2C, D).

To further understand the functional effect of the R599C mutation we employed affinity purification combined with mass spectrometry (AP-MS) to examine and understand the stable interactions and proximity-dependent biotin identification (BioID) to identify transient and close-range interactions. A comparative analysis of the high confidence interactors (HCI) between the WT and R599C receptor revealed a significant difference in the interacting partners (Supplemental Table 1) and an enrichment of signaling pathways. Both loss of interacting partners (eg. PLCG1, ELMO2), and novel interactor partners (eg. PDIA4 and SSRP1) were observed in the R599C mutant. In addition, the R599C mutation resulted in the reduction of interaction partners overall. GO analysis of the results indicated that the R599C mutated receptor had a reduction of interaction partners specifically in the ERBB2 signaling pathway (Supplemental Figure 1).

### ERBB2 R599C receptor interacts with proteins in endoplasmic reticulum and mitochondria

To gain a deeper understanding of the potential compartment-specific localization of ERBB2, we utilized our developed MS-microscopy system with the BioID data (44). This system combines quantitative interactome profiling with microscopy techniques to accurately map the cellular distribution of the ERBB2 protein. The ERBB2 WT receptor was identified to be localized mainly at the plasma membrane, as expected. Interestingly, the ERBB2 R599C receptor was predicted to localize mostly to mitochondria and ER based on its interaction partners (Figure 2E).

To confirm these findings in patients, we examined the plasma membrane expression of ERBB2 in hiPS-CMs from a patient and from healthy controls. We acquired fibroblasts from the index patient of the Family 1, created hiPSCs and differentiated them into cardiomyocytes. First, we ascertained that the variant allele is expressed in the patient cells. Digital PCR results showed that in both hiPSCs and hiPSC-CMs the variant allele accounted for approx. 50% of the *ERBB2* gene expression (Figure 2F). HiPS-CMs from the index patient and from two healthy individuals (HEL24.3, HEL47.2) were then stained with an ERBB2 antibody that binds to the extracellular part of the receptor and the cells were analysed with flow cytometry. Indeed, the results were consistent with the MS microscopy findings showing that the patient hiPS-CMs had approximately half of the staining intensity of the control cells on the cell membrane, further supporting the hypothesis that the mutant protein localizes mostly intracellularly (Figure 2G).

### Zebrafish embryos expressing the ERBB2 R599C receptor have reduced cardiac contractility and increased cardiac wall thickness

Next, we examined the functional role of the ERBB2 R599C receptor in the heart. Zebrafish embryos were injected with a plasmid containing either *ERBB2* WT gene or *ERBB2* R599C together with mCherry to enable visualization of the injected cells (Supplemental Figure 2). Two control groups, one with an empty vector and one with non-injected embryos, were included. There was no difference in the fractional shortening between *ERBB2* WT injected, vector injected or non-injected hearts (Figure 3A), showing that the plasmid injection itself or human ERBB2 WT overexpression did not affect cardiac function. However, zebrafish embryos injected with the *ERBB2* R599C plasmid demonstrated significantly lower fractional shortening compared to the *ERBB2* WT injected embryos and the two control groups (Figure 3A), indicating compromised function of the heart caused by the R599C variant. In searching for underlying physical defects in the heart, we discovered that the cardiac wall was significantly thicker in the ERBB2 R599C embryos, measured from brightfield movies (Figure 3B). By using whole-mount actin staining we could determine that specifically the myocardium was thicker in the ERBB2 R599C embryos compared to ERBB2 WT embryos (Figure 3B). To determine whether the thickening of the myocardium was a result from hyperplasia or hypertrophy of the cells, the number of nuclei were counted, and the area of the cells was determined from the mCherry positive cells. The results showed no difference in the number of nuclei, indicating no hyperplasia (Figure 3C). mCherry staining, in turn, revealed increased surface area of the mutant cells (Figure 3D, E), confirming that the thickening of the myocardium was due to hypertrophy of the cardiomyocytes.

**Figure 3:**
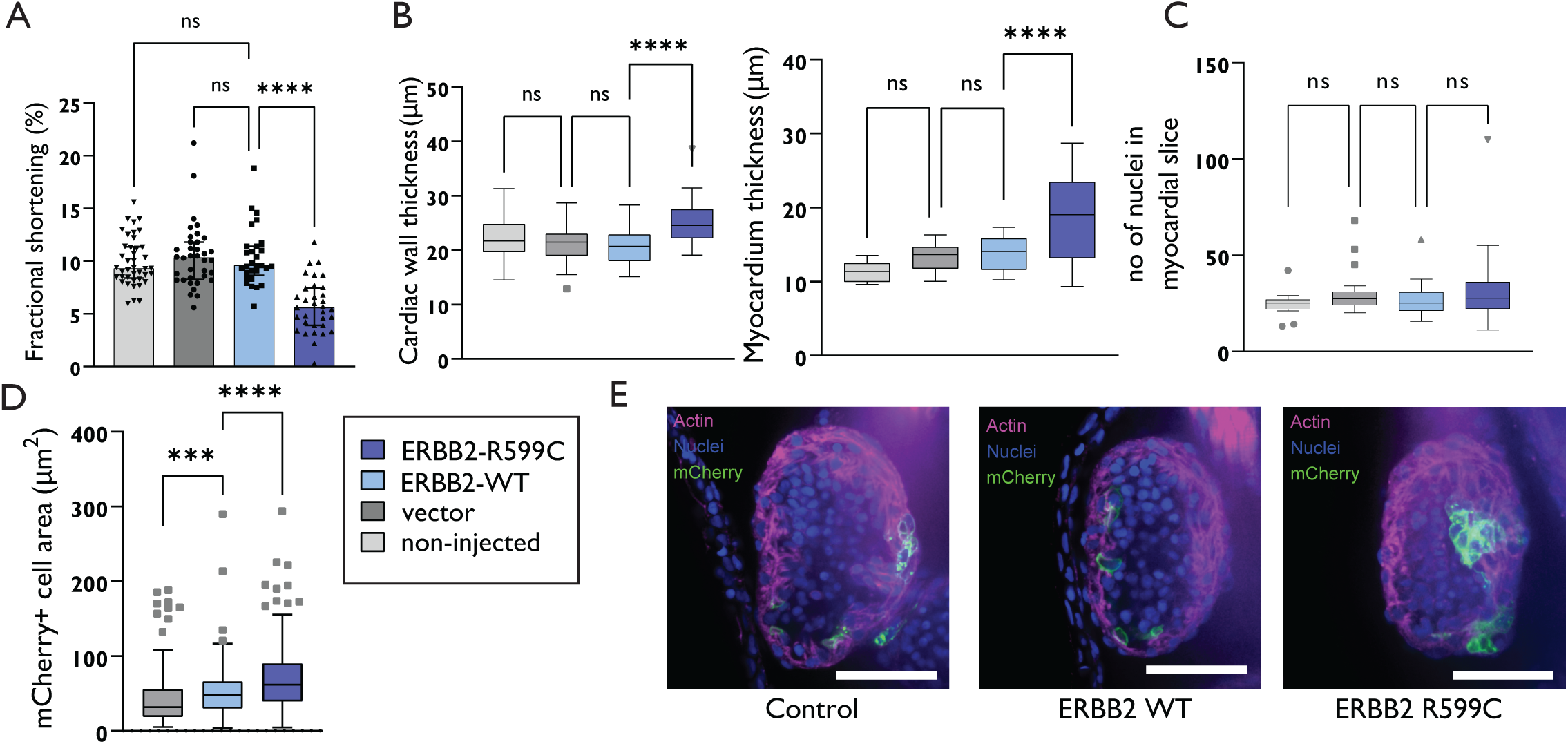
A) Fractional shortening of the heart of the zebrafish embryos injected with the ERBB2 R599C plasmid, ERBB2 WT plasmid, empty vector, and no injection indicate decreased pump function in the zebrafish embryos injected with the ERBB2 R599C plasmid. B) Increased cardiac wall thickness as measured from brightfield videos and increased myocardium thickness measured by phalloidin staining, is seen in the zebrafish embryos injected with the ERBB2 R599C plasmid. C) No differences in the number of nuclei within the four experimental groups was seen. D) Cell area measured by mCherry staining in each group indicated slightly enlarged cells in the zebrafish embryo injected with the ERBB2 WT and ERBB2 R599C plasmid. E) Example images of phalloidin staining of the heart. Scalebar 50 μm.

### ERBB2 R599C patient hiPSC-derived cardiomyocytes and endothelial cells have aberrant expression of genes related to cardiovascular system development

HiPS cells derived from the LVOTO patient with the *ERBB2* R599C variant and from four healthy controls (HEL24.3, HEL47.2, HEL46.11, K1) were differentiated into cardiomyocytes and single-cell RNA sequencing (scRNA-seq) was performed when the cells were approximately 35 days old. Six clusters were identified in the UMAP (Figure 4A), with all clusters expressing cardiomyocyte-specific genes (Figure 4B), demonstrating efficient differentiation and selection. Pathway analysis revealed altered expression of genes related to heart development, muscle contraction, cardiac muscle hypertrophy, and stress responses in the LVOTO patient cells (Figure 4C, Supplemental Table 2). Compared to the healthy cells the LVOTO patient cells had significantly lower expression of cardiac genes such as *MYH6, NPPA* and *NPPB* and higher expression of *MYH7* (Figure 4D, Supplemental Table 2). Finally, the LVOTO patient cardiomyocytes had lower expression of many genes related to oxidative stress (Figure 4E, Supplemental Table 2).

**Figure 4:**
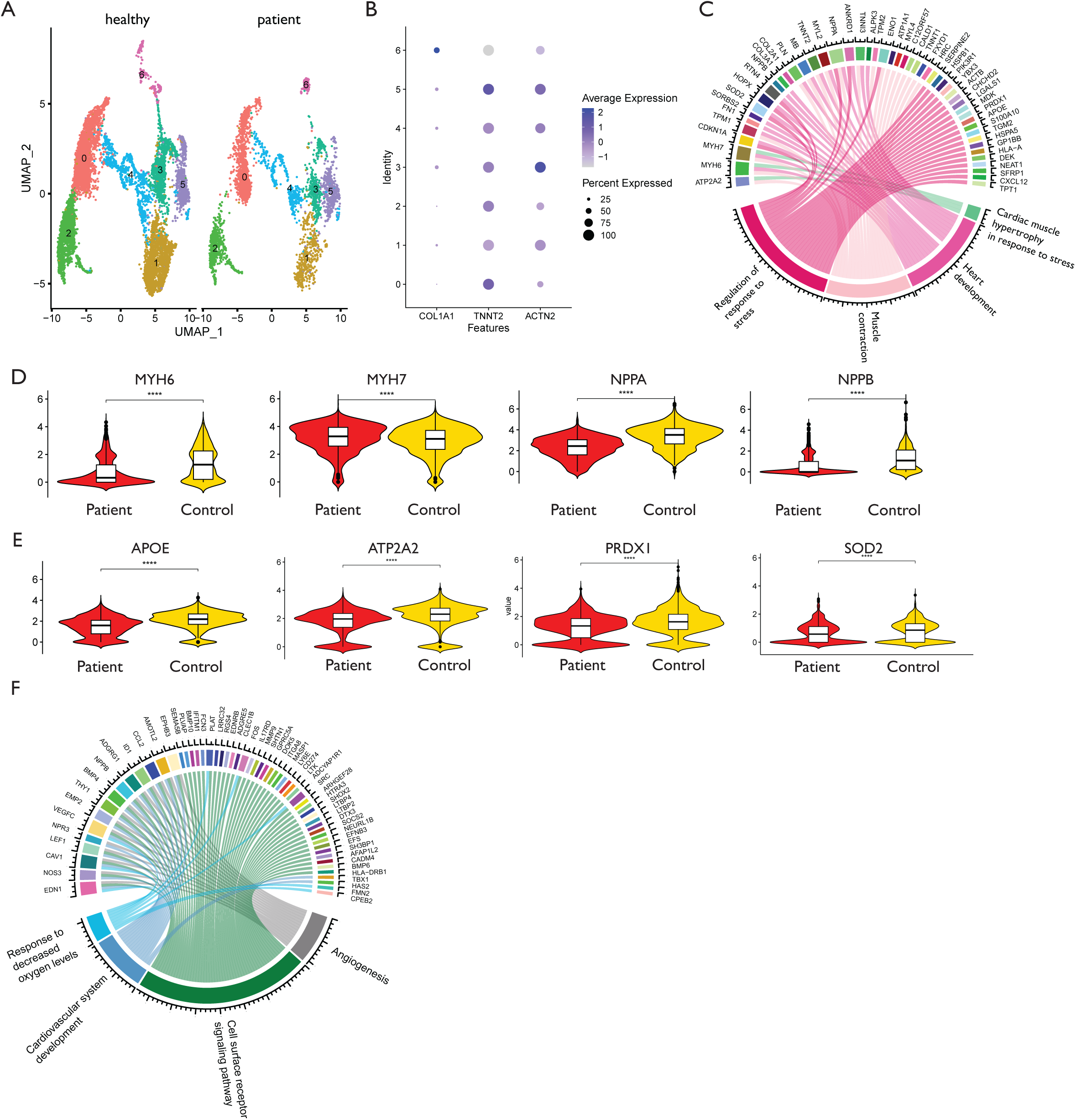
A) Uniform Manifold Approximation and Projection (UMAP) of patient and healthy control hiPS-CMs (control cells combined from four healthy individuals). B) Dot plot shows clusterwise expression of cardiomyocyte marker genes TNNT2 and ACTN2 and the fibroblast marker COL1A1. C) Circos plot of pathways that are differentially expressed in hiPS-CMs of the patient with the ERBB2 R599C variant according to gene ontology GO analysis. Violin plots presenting the expression levels of D) heart development and E) oxidative stress related genes in patient and control cellls. F) Circos plot of pathways that are differentially expressed in patient hiPS-ECs as compared to those of healthy controls.

As proper heart development involves intimate crosstalk between endothelial/endocardial cells and cardiomyocytes, we studied the effect of the *ERBB2* R599C variant on endothelial cells. The LVOTO patient hiPSCs and cells from three healthy individuals (HEL47.2, HEL24.3, K1) were differentiated into ECs, and two independent differentiation batches were analysed with whole-genome RNA sequencing. There were 198 repeatedly differentially expressed genes between the LVOTO patient and control hiPS-ECs (Supplemental Table 2). The LVOTO patient hiPS-EC samples clustered together away from the control samples, demonstrating reproducibility between the two independent differentiations (Supplemental Figure 3). Repressed expression in genes related to cardiovascular system development, response to decreased oxygen levels, programmed cell death, cell surface signaling pathway and angiogenesis was observed, including genes such as *NPPB, NOS3, SRC* and *BMP10* (Figure 4F, Supplemental Table 2).

## Discussion

We have identified a rare *ERBB2* missense variant R599C in three unrelated families with LVOTO defects. The variant caused protein mislocalization, striking differences in binding partners, altered transcriptomics in patient derived hiPS-cardiomyocytes and -endothelial cells, and resulted in developmental defects in zebrafish embryo hearts, providing functional evidence suggesting *ERBB2* as a novel disease gene for CHD.

The *ERBB2* (Erb-B2 Receptor Tyrosine Kinase) gene encodes one of four members of the epidermal growth factor (EGF) receptor family of receptor tyrosine kinases. ErbB receptor dimerization by neuregulin leads to tyrosine kinase activation, which plays a vital role in embryogenesis (27, 45). *ERBB2* has not been previously associated with CHDs in humans even though its essential role in cardiac morphogenesis, especially in cardiac wall trabeculation, is well established in animal models (27, 46–48). In addition, the importance of ERBB2 signaling in the adult heart has been demonstrated when trastuzumab, an ErbB2 monoclonal antibody used in breast cancer therapy, was shown to associate with increased left ventricular dysfunction when used in combination with anthracyclines (49).

Unlike other EGF receptors, ERBB2 has no ligand binding domain of its own and therefore cannot bind growth factors. However, it does bind tightly to other ligand-binding EGF receptor family members to form heterodimers, enhancing kinase-mediated activation of downstream signaling pathways via receptor phosphorylation (50). Our results show that the change of arginine to cysteine in the ERBB2 R599C mutant does not affect the phosphorylation capability of the receptor. As the phenotype could not be explained by defective phosphorylation, we explored the interactions of the ERBB2 R599C mutant receptor with other proteins. The results revealed that the mutant receptor has overall fewer and different binding partners compared to the wild type receptor. According to the MS microscopy, the ERBB2 R599C mutant receptors binding partners localize to the mitochondria and ER, and in line with this result, the patient hiPS-CMs had roughly 50% reduced expression of ERBB2 on the plasma membrane compared to healthy cells. These results indicate that the R599C variant leads to mislocalization of the protein intracellularly. The change in binding partners may be due to the free cysteine allowing the formation of an extra cysteine bond, or due to changes in the folding of the protein. The extra cysteine may also contribute to changes in protein-protein interaction via non-covalent interactions involving the sulfur atoms of the cysteine residue (51). Although the cytosol as a reducing environment is not conducive to formation of disulfide bonds (52, 53), disulfide bonds can be formed in the ER and mitochondria (54). It has been reported that ERBB2 also localizes within the mitochondria of both cancer cells and other diseases (55).

GO results from the protein-protein interaction assay indicate overall reduction of ERBB2 and receptor tyrosine kinase signaling in cells with the ERBB2 R599C receptor. The GO results also indicate reduced interaction with the PI3K-PKB/Akt pathway, which is one of the downstream signaling pathways of the ERBB2 receptor (56). Thus, as the mutant receptor phosphorylation was not deficient, it is likely that protein mislocalization and/or altered binding partners cause the impaired ERBB2 signaling. Defective ERBB2 signaling may lead to e.g. reduced proliferation of cells, as proliferation related GO terms are only found in the WT ERBB2 interactions.

Many of the PI3K-PKB/Akt pathway proteins with reduced interaction with the ERBB2 R599C receptor have important roles in heart development or function. PRKCI is required for heart trabeculation in mice (57), PIK3R2 regulates heart size and hypertrophy in mice (58), PIK3CB promotes CM proliferation and survival in neonatal rat CMs (59) and PRKCA regulates heart contractility in mice (60). In addition, the ERBB2 R599C receptor had reduced interaction with proteins such as SOS1, PTPN11, CBL, which have been associated with syndromic CHD (61, 62) (63) and MICOS13, DTNA, and EMC1 which have been associated with nonsyndromic CHD (64) (65, 66). In addition, the mutant receptor has lost interaction with FRS2, and Frs2alpha is required for outflow tract morphogenesis (67). Loss of interaction with these proteins that are implicated to be important to heart development may contribute to observed heart defects in the individuals with the mutant receptor.

To study the functional effect of the ERBB2 R599C mutant receptor in vivo, we used zebrafish, as it is a commonly used model for heart development and regeneration (68, 69). Interestingly, expression of the mutant human ERBB2 in zebrafish embryo hearts led to cardiomyocyte hypertrophy, increased cardiac wall thickness, and impaired fractional shortening of the heart demonstrating that the presence of the mutant receptor induces functional defects during heart development. Animal models of HLHS have shown that intrinsic myocardial defects are associated with HLHS (70, 71), and a recent study in zebrafish demonstrated that *rbfox* mediated reduction in pump function led to compromised development of the valves and aorta (71). Thus, our findings recapitulate this predisposition for outflow tract obstruction tract development.

When modeling HLHS at the cellular level, transcriptomic and functional studies on HLHS-patient derived hiPS-CMs show impaired differentiation (72, 73), less organized sarcomere structure (72–74), and reduced contractility (75) that likely are associated with the compromised pump function seen in animal models of HLHS. Our results were in line with these studies. The transcriptomic analyses of ERBB2 R599C hiPSC-CMs provide evidence that several genes and pathways related to heart and cardiovascular system development were affected in the patient cells. Among the downregulated were two genes encoding sarcomere proteins, MYH6, which has been associated with HLHS (74), and TNNI3, which has been associated with dilated, hypertrophic and familial restrictive cardiomyopathy (76–78). Reduced expression of TNNI3 in HLHS hiPS-CMs has also been demonstrated in a previous study (72). In contrast, gene expression of the sarcomere protein MYH7 was increased in the ERBB2 R599C hiPS-CMs, and interestingly, increased MYH7 expression has been shown in atrial and ventricular tissues of HLHS subjects, and in hiPS-CMs derived from HLHS patients (74). Moreover, similar increases of TNNT2 and MYL2 expression in HLHS hiPS-CMs observed in our study have been documented previously (74).

Intrinsic endocardial defects contributing to abnormal valve formation have been associated as a potential mechanism underlying HLHS (79), and loss of Erbb2 in coronary endothelial cells during development has been associated with improper patterning of coronary vasculature (80), demonstrating its important role in endothelial cells in cardiac development. Intimate crosstalk between ECs and CMs is essential during cardiac development. Therefore, we also studied the patient hiPSC-derived ECs using RNA-seq in comparison with cells from healthy controls.

The transcriptomic analyses of patient hiPS-ECs provide evidence that several genes and pathways related to heart and cardiovascular system development were affected in the patient cells. Expression of BMP10 was downregulated in ERBB2 R599C hiPS-ECs. Research has highlighted the significance of BMP10 in preserving the expression of genes crucial for cardiac development, including NKX2.5, which has been identified as being associated with HLHS (81, 82). Additionally, BMP10-null mice have been shown to have defects in cardiomyocyte proliferation and trabeculation (82). These findings further support the theory that ECs have a role in the development of HLHS.

Abnormal response to oxidative stress is associated with congenital heart defects (83–85) (86). A recent study showed reduced mitochondrial respiration and oxidative metabolism in HLHS hiPS-CMs compared to healthy controls potentially contributing to reduced contractility (75). Several oxidative stress and metabolic genes were downregulated both in patient hiPS-ECs and -CMs. The role of ERBB2 in endothelial ischemic conditions has been demonstrated in a mice model, where ERBB2-signaling was shown to facilitate the recruitment of SRC. This recruitment subsequently activated NOS3 through Nrg1, leading to enhanced myocardial perfusion by elevating nitric oxide production and promoting vasorelaxation (87). Interestingly, a decrease in expression was observed for both SRC and NOS3 in ERBB2 R599C hiPS-ECs. NOS3-deficiency has demonstrated an association with a bicuspid aortic valve in mice. This connection possibly arises from the role of the valvular endothelium in fine-tuning the developmental process through mechanisms like shear stress and other luminal events (88). Moreover, reduced expression in metabolic genes including ENO1, SOD2 and SOD3 was observed in the ERBB2 R599C hiPS-CMs. These genes have been shown to provide protection under hypoxic and oxidative stress (75, 89, 90).

ERBB2 variants have not previously been associated with CHD in human, but one study has reported copy number variants including an *ERBB2* gene duplication in a patient with total anomalous pulmonary venous return and in a patient with Tetralogy of Fallot (91). However, no further experiments were performed to validate the findings or to further investigate the potential role of ERBB2 as a causal gene.

While most persons with the ERBB2 R559C variant had CHD, there were two persons with this variant who had normal echocardiographic findings and one person without reported cardiac symptoms but in whom no echocardiographic data were available. This is not unexpected, as reduced penetrance has been associated with monogenic CHD in multiple studies. For example, *NOTCH1* haploinsufficiency is a well-known cause of CHD with a penetrance reported to be about 75% (22, 24). In addition, this study demonstrated well the variable expressivity associated with CHD (22, 24), as the cardiac abnormalities ranged from BAV and CoA to HLHS and Shone’s complex. Of note, the phenotype severity seemed to worsen with consecutive generations. This could be due to accumulation of additional predisposing variants, or environmental triggers. However, this can also depend on case selection, as our study cohort comprised on average more severe pediatric cases.

Previous research shows that total ERBB2 deletion in mice leads to lethal cardiac malformations early during development (27), while mice with conditional knockout of ERBB2 in ventricular myocytes show normal cardiac morphogenesis at birth and survive to adulthood (92). However, in adulthood these mice show features of dilated cardiomyopathy including biventricular enlargement, decreased cardiac wall thickness, and decreased fractional shortening (92). Thus, it seems clear that the role of ERBB2 signaling in the adult heart is different from its role during embryogenesis, and that during development, ERBB2 signaling is essential in several different cell types. Nevertheless, the recent observation that reduced ventricular contractility led to compromised development of the valves and aorta in zebrafish (71) supports the hypothesis that intrinsic myocardial defects, possibly mediated by additional cell types such as endocardial or endothelial cells, are causal for the phenotype observed in our affected study subjects.

Taken together, our results provide strong evidence for ERBB2 as a new disease gene for LVOTO defects. The ERBB2 R599C variant was present in all affected members of three unrelated families with CHD in multiple generations and was only found in two individuals in gnomAD. Functional analysis of the mutant receptor demonstrated dramatic changes in the binding partners and cellular localization of the receptor. Finally, expression of the mutant allele caused compromised heart function in the zebrafish. Further analysis on the exact mechanisms of the mutant protein at the cellular level will provide more information on the pathogenic events leading to CHD during cardiac development.

---

Supplemental Figure 1. Gene ontology analysis of ERBB2 WT/ERBB2 R599C interaction partners. A) GO Biological process, B) KEGG pathway analysis, C) Reactome Pathway.

Supplemental Figure 2: A) Heatmap showing the patient and control samples.

Supplemental Figure 3: A) Plasmid map of the vector used for zebrafish embryo transfection. Supplemental Table 1. ERBB2 WT/ERBB2 R599C interaction partners.

Supplemental Table 2. CM scRNA-seq and EC RNA-seq differentially expressed genes.

## Data Availability

All data produced are available online at GEO and MassIVE (MSV000093153).

https://massive.ucsd.edu/ProteoSAFe/dataset.jsp?task=1fbfd8a81ba14008af86bf3280f7506f

## Acknowledgements

We thank Ilse Paetau for her help in the cell culture, laboratory experiments and administrative support. Exome sequencing and data analysis were provided by the University of Washington Center for Rare Disease Research (UW-CRDR). The content is solely the responsibility of the authors and does not necessarily represent the official views of the National Institutes of Health. We thank Zebrafish Core and Cell Imaging Core (both Turku Bioscience Centre, University of Turku and Åbo Akademi University, and supported by Biocenter Finland) for instrumentation and services. We thank Professor Timo Otonkoski and Docent Ras Trokovic at Biomedicum Stem Cell Center for providing us with three control hiPS-cell lines and Professor Anu Suomalainen-Wartiovaara for the gift of the hiPSC line K1. Institute of Molecular Medicine Finland (FIMM) is acknowledged for providing the scRNA-seq service and Biomedicum Functional Genomics Unit (FuGU) for providing the bulk RNA-seq service. The flow cytometry analysis was performed at the HiLife Flow Cytometry Unit, University of Helsinki.

## Data Availability Statement

The sequencing data underlying this article will be available in GEO under accession numbers GSE247858 and GSE194103. The mass spectrometry data will be available at MassIVE with dataset ID MSV000093153 (https://massive.ucsd.edu/ProteoSAFe/dataset.jsp?task=1fbfd8a81ba14008af86bf3280f7506f).

