## Supplementary figures and images for "ERBB2 R599C variant is associated with left ventricular outflow tract obstruction defects in human"

### Supplemental Figure 1

Supplemental Figure 1.

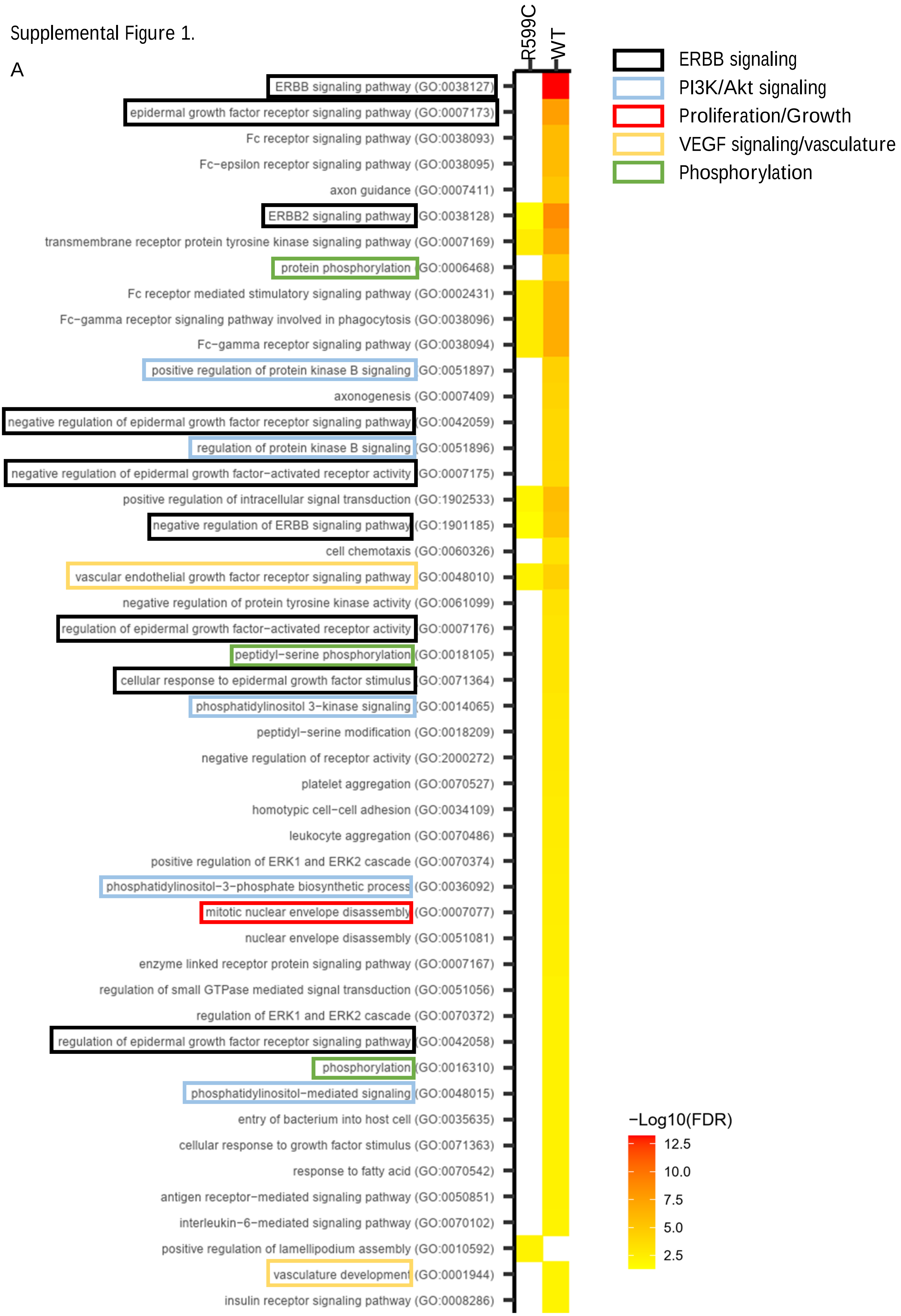

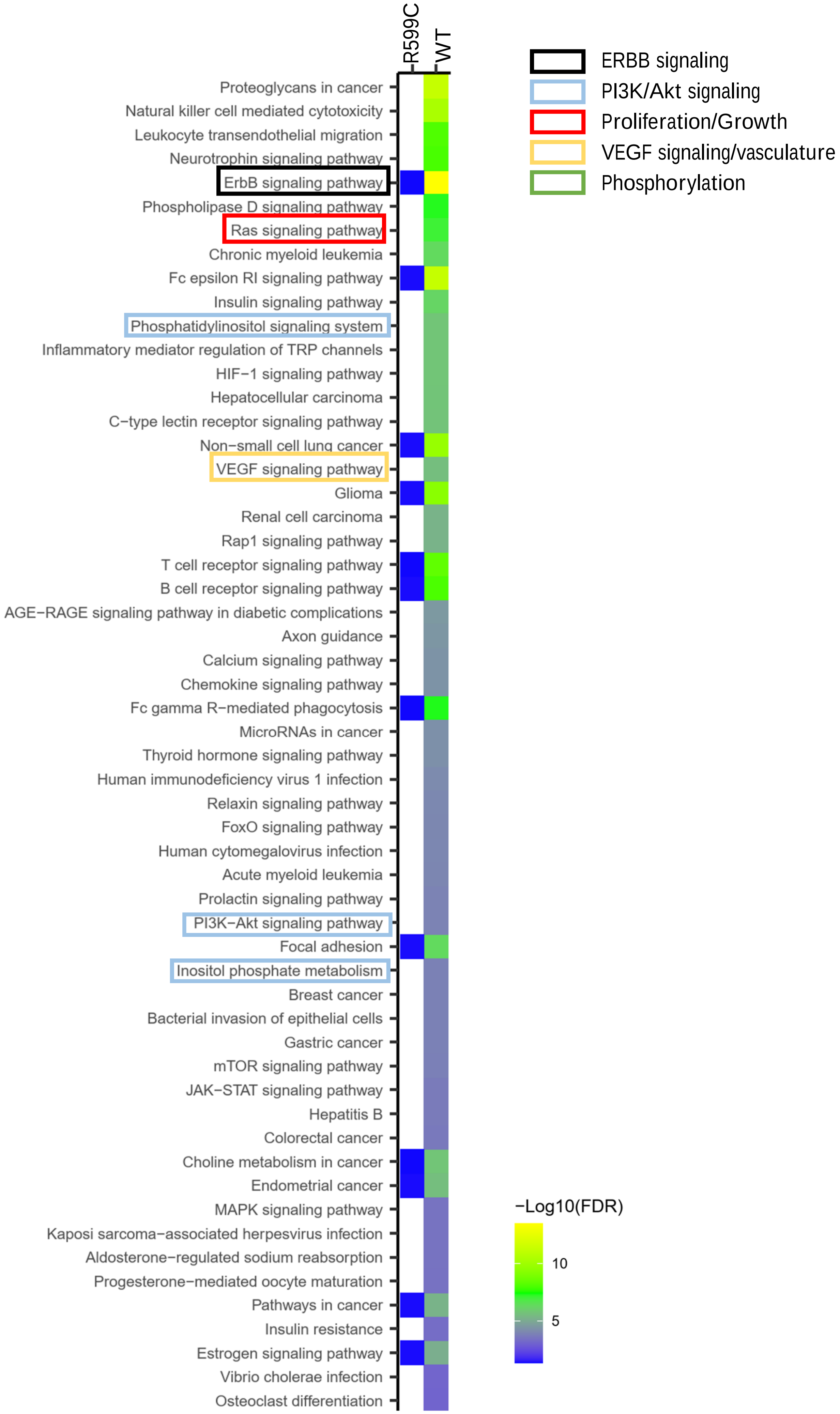

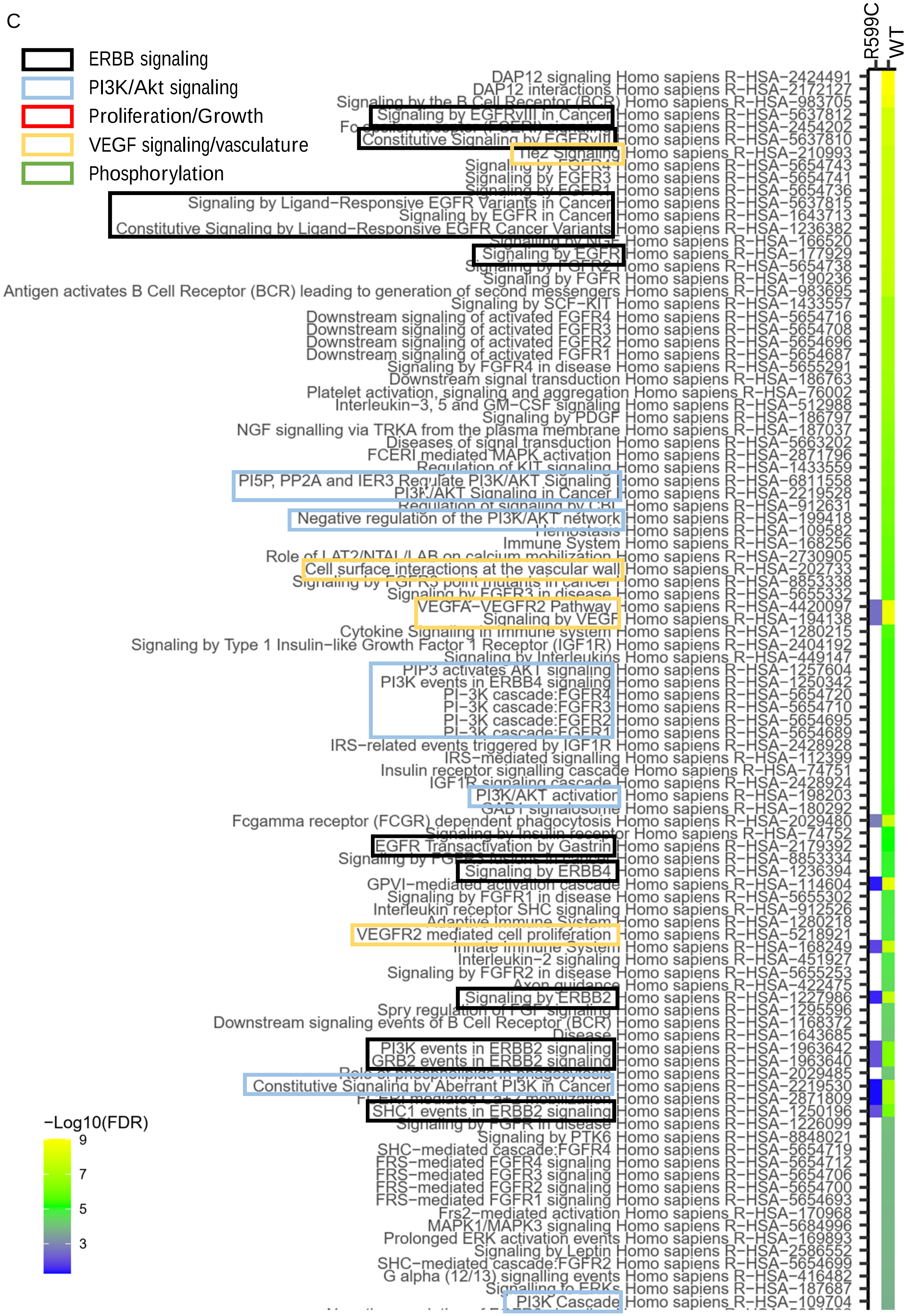

### Supplemental Figure 2

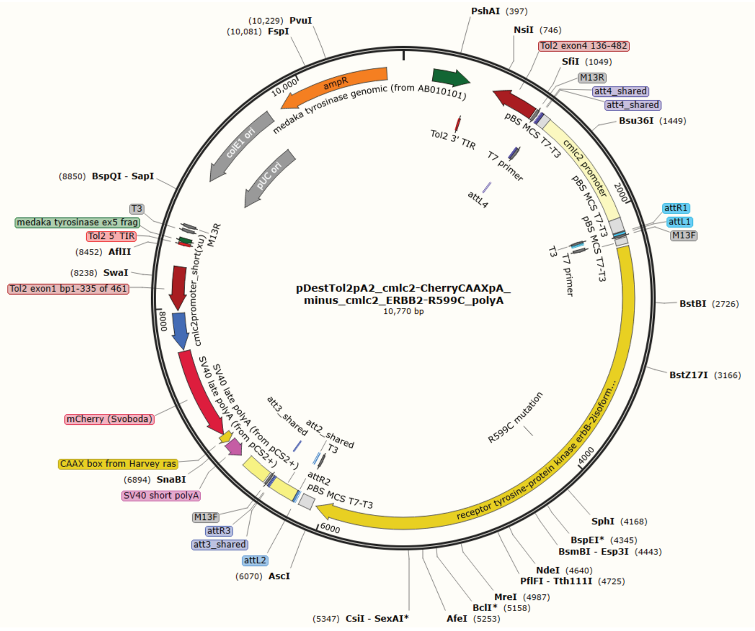

### Supplemental Figure 3

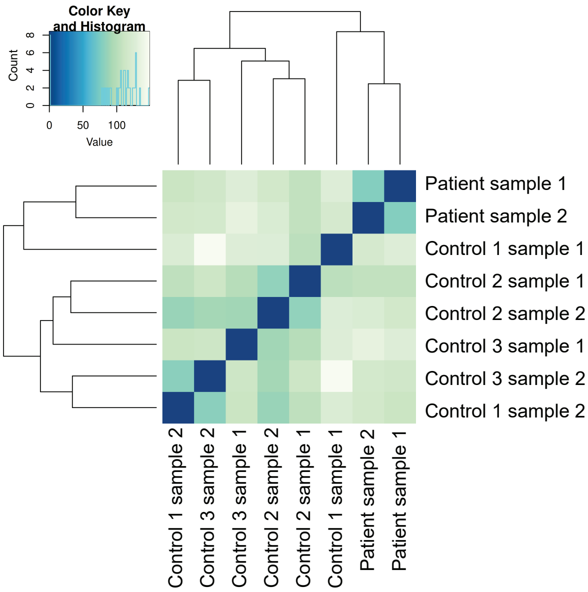
