## Supplemental Methods for "ERBB2 R599C variant is associated with left ventricular outflow tract obstruction defects in human"

*Exome sequencing*

**UW-CMG cohort.** In brief, library capture was performed with Roche/Nimblegen SeqCap EZ v2.0, with 75-base pair paired-end sequencing on the HiSeq2500/4000 instrument (RTA 1.18.34/RTA 2.5.2). BAM files were aligned to a human reference (hg19hs37d5) using BWA-MEM (Burrows-Wheeler Aligner; v0.7.10) (1). Read-pairs not mapping within ± 2 standard deviations of the average library size (~150 ± 15 bp for exomes) were removed. RTG-core version 3.3.2 was applied to the raw exome sequence data for mapping, pedigree-aware variant calling, and genotype filtration (Real Time Genomics Inc., Hamilton, New Zealand) (2).

**BPG cohort.** DNA was fragmented using Covaris (Covaris, Inc. Woburn, MA). The sequencing library preparation was done using NEBNext Ultra II library kit (New England Biolabs, Ipswich, MA, USA) and indexed adapters (Integrated DNA Technologies, Coralville, IA, USA). WES capture was performed using IDT xGen Exome Research Panel assay (Integrated DNA Technologies, Coralville, IA, USA) with spiked-in custom designed clinical content. Sequencing was performed using the Illumina NovaSeq 6000 (Illumina, San Diego, CA, USA).  Raw sequencing data was demultiplexed using bcl2fastq software and aligned to GRCh37 (hg19) build of the human reference genome using BWA software with mem algorithm (1). SNV and Indel variants were called using Sentieon Genomics (release 201711).

*p-ERBB2/ERBB2 Western blot, disulfide bridge Western blot*

hErbb2 was amplified in three parts; 5'-end was amplified from cloned cDNA from buccal swap using TAGACGAAGCTTGGGCTGCAGGTCGACTCTAGAGCCACCATGGAGCTGGCGGCC

TTG and GATCTCTGTGAGGCTTCGAAGC, 3'-end was amplified in two parts from ATCC cDNA-clone pCER204, which lacks the first 300 bp, using CTGCGGGAGCTGCAGCTTC and **GGTTTCACACCGCTGGGGCAGCAGGCCACGCAGAAGGGAG**, **TCTGCGTGGCCT GCTGCCCCAGCGGTGTGAAA**, ATCGATAAGCTTGATATCGAATTCGGCGCGCCTC ACACTGGCACGTCCAG. Primers marked in bold carried R599C-mutation. The fragments had primer-created homology with each other and the vector and were assembled with FUW-XbaI-AscI and are described as FUW-hErbB2-WT or FUW-hErbB2-R599C from here on. Cos-7 cells were cultured in DMEM supplemented with 10% FBS, l-glutamine and penicillin-streptomycin. 140,000 Cos7 cells per well were plated in 6-well plates. They were simultaneously reverse transfected using 1ug of plasmids (sham, FUW-hErbB2 -WT or FUW-hErbB2-R599C), 3µl of Fugene HD and 50µl of Optimem I. Cells were incubated at 37C° cell culture incubator overnight. This was followed by two washes of cells with PBS, and the addition of 2ml of serum-free DMEM in each well. Cells were serum starved for 20h and then cells were lysed with 100µl of 1x sample buffer (50 mM Tris-HCl pH 6.5, 2% SDS, 6.25% glycerol, 1.5 mM bromophenol blue, 0.2 M dithiothreitol), boiled 5 min at 95C° and stored at -20C° until analysed. For analysis of disulfide bridges, the cells were not serum-starved and were lysed into TXLB lysis buffer (50 mM Hepes, 1% Triton X-100, 0.5% sodium deoxycholate, 0.1% SDS, 0.5 mM EDTA, 50 mM NaF, 10 mM Na3VO4, supplemented with protease inhibitor cocktail (cOmplete Mini, EDTA-free, Roche)). The samples were boiled in sample buffer with or without dithiotreitol.

*RNA extraction, cDNA synthesis and dPCR*

NucleoSpin RNA kit (Macherey-Nagel, 740955.250) was used for RNA extraction. cDNA synthesis was performed using High-Capacity cDNA Reverse Transcription Kit (Thermo Fisher, 4368814). A custom assay was designed for identification of WT and mutant allele expression of *ERBB2* gene (dPCR LNA Mutation Assay, Qiagen, 250200). The assay contains primers, a HEX-conjugated probe that detects the WT allele and a FAM-conjugated probe that detects the mutant allele. QIAcuity Probe PCR Kit (Qiagen, 250101) was used for the reaction performed with QIAcuity One (Qiagen). cDNA samples from HLHS patient and healthy hiPSCs and hiPS-CMs were used.

*Protein-protein interactions with AP and Bio-ID*

Entry clone containing ERBB2 WT was obtained from the human ORFeome collection v9.1 (Center for Cancer Systems Biology, Dana-Farber Cancer Institute). Using PCR amplification ERBB2 R599C entry clone was generated by designing mutation inducing primers. ERBB2 WT and R599C constructs were fused with MAC-Tag-C (Addgene, Plasmid #108077) destination vector using Gateway cloning techniques. Flp-In™ 293 T-REx cell line (Invitrogen, R78007) was used to generate isogenic and inducible stable wild type and mutant expressing cell lines as in Liu et al. 2020 (3). Each stable cell line was expanded to 80% confluence in 15 × 150 mm cell culture plates (CELLSTAR, Greiner). Five plates were used for each replicate, in which 1 μg/ml tetracyclin and 50 μM biotin was added for 24 h before harvesting. Pervanadate treatment was performed at a concentration of 100 µM for 15 min prior to harvesting. Cells from each replicate (n=3) were pelleted and snap-frozen and stored at −80 °C.

Cells were lysed in ice‐cold lysis buffer supplemented with 0.5 mM PMSF and protease inhibitors. For AP‐MS, samples were lysed in 3 ml of ice‐cold lysis buffer (0.5% IGEPAL, 50 mM HEPES (pH 8.0), 150 mM NaCl, 50 mM NaF, 1.5 mM NaVO3, 5 mM EDTA, 0.5 mM PMSF, and protease inhibitors (Sigma‐Aldrich)). For BioID‐MS approach, cell pellets were thawed in 3 ml of ice‐cold lysis buffer (0.5% IGEPAL, 50 mM HEPES (pH 8.0), 150 mM NaCl, 50 mM NaF, 1.5 mM NaVO3, 5 mM EDTA, 0.1% SDS, 0.5 mM PMSF, and protease inhibitors), and lysates were sonicated and treated with Benzonase® Nuclease (Santa Cruz Biotechnology, sc‐202391).

Cleared lysate was obtained by centrifugation, and the lysate was subjected to a one‐step purification via Strep‐Tactin® Sepharose® resin (IBA). The purified protein complexes were reduced, alkylated, and digested to peptides for MS analysis (3).

The desalted samples were analysed using the Evosep One liquid chromatography system coupled to a hybrid trapped ion mobility quadrupole TOF mass spectrometer (Bruker timsTOF Pro) via a CaptiveSpray nano-electrospray ion source. An 8 cm × 150 µm column with 1.5 µm C18 beads (EV1109, Evosep) was used for peptide separation with the 60 samples per day methods (21 min gradient time). Mobile phases A and B were 0.1 % formic acid in water and 0.1 % formic acid in acetonitrile, respectively. The MS analysis was performed in the positive-ion mode using data-dependent acquisition (DDA) in PASEF mode (4) with DDA-PASEF-short_gradient_0.5s-cycletime -method.

Raw data (.d) were processed with FragPipe v17.1 utilizing MSFragger (5) against reviewed human entries of the UniProtKB database (downloaded 8.3.2022). Carbamidomethylation of cysteine residues was used as static modification. Aminoterminal acetylation and oxidation of methionine were used as the dynamic modification. Biotinylation of lysine and N-termini were set as variable modifications. Trypsin was selected as enzyme, and maximum of two missed cleavages were allowed. Both instrument and label-free quantification parameters were left to default settings. Final results from these steps are Spectral Counts (SC) values from peptides with FDR < 0.01 from Philosopher.

Significance Analysis of INTeractome (SAINT) -express version 3.6.3 (6) and Contaminant Repository for Affinity Purification (CRAPome, (7)) were used to discover statistically significant interactions from the data. Final results represent proteins with a BFDR < 0.01, and in less than 20% of Crapome database experiments except in cases where AvgSpec is three times higher than AvgSpec in Crapome experiments. Data visualization was performed with Prohits-viz.org and proteomics.fi webtools.

*Functional and pathway enrichment*

All the high confidence interactors obtained from AP-MS and BioID were subjected to KEGG database (<https://www.genome.jp/kegg/>) (8), Reactome pathway-based enrichment (<https://reactome.org/>) (9) and David Bioinformatics ([https://david-d.ncifcrf.gov](https://david-d.ncifcrf.gov/)) (10) to obtain gene ontology (GO) enriched terms with *P* value<0.05.

*Plasmid cloning*

Zebrafish Tol2 transgenesis plasmids harboring zebrafish cmcl2-promoter (11) to drive expression in zebrafish myocardium were cloned by utilizing Tol2kit protocols (12). First, in vitro synthesized DNA (gBlock, Integrated DNA Technologies) cmcl2:mCherry-CAAX:pA fragment carrying and pDestTol2A2 plasmid were digested with BglII, plasmid dephosphorylated with Antarctic Phosphatase (NEB) and products ligated with T4 DNA ligase (NEB). Ligation reactions were transformed into ccdB resistant competent bacteria (Invitrogen) and selected on Ampicillin+Chloramphenicol plates. Resulting clones were grown as minipreps, DNA isolated with Macherey-Nagel kit and screened with BglII and BstXI digestions to yield plasmid pDestTol2pA2-cmlc2CherryCAAX. Next, the WT and mutant ERBB2 inserts were excised from pFUW plasmids using SalI-HindIII digestion and ligated to SalI-HindIII digested pME-MCS plasmid. The ligation reactions were transformed into DH10b (NEB) competent bacteria and selected on kanamycin plates. Final transgenesis vectors were generated in three multisite Gateway LR recombination reactions using LR Clonase II (Invitrogen) and following plasmids: 1) pDestTol2pA2-cmlc2CherryCAAX, p5E-cmcl2, pME-MCS and p3E-polyA; 2) pDestTol2pA2-cmlc2CherryCAAX, p5E-cmcl2, pME-ERBB2-WT, p3EpolyA; 3) pDestTol2pA2-cmlc2CherryCAAX, p5E-cmlc2, pME-ERBB2 R599C, p3E-polyA. Recombinants were transformed into DH10beta bacteria, selected on ampicillin plates, grown as minipreps and identified using diagnostic KpnI digestion. Final clones were sent to full plasmid sequencing performed by e-Zyvec (Loos, France). Plasmid p5E-cmlc2 (11) was a gift from Dr. Xiaolei Xu (Mayo Clinic) and plasmids pME-MCS, p3E-polyA and pDestTol2pA2 were gifts from Dr. Chi-Bin Chien (University of Utah Medical Center) and described in (12).

*Zebrafish stainings*

Zebrafish embryos were fixed in 4% formaldehyde in PBS supplemented with 0.2% Tween-20. Five groups of 10 embryos, cmlc2:MCS, cmlc2:ERBB2-WT, cmlc2:ERBB2-R599C and two uninjected, were washed twice with PBSTx (PBS+0,2% Triton X-100). 1:10 trypsin solution in PBSTx was added and embryos were incubated for 2 min. Embryos were washed again twice with PBSTx. After that they were permeabilized with 2% Triton X-100 in PBS for 2 h and washed twice with PBSTx. Embryos were incubated for 2 h with blocking solution 5 % FBS in PBSTx. Then 1.25µ/ml Alexa-647-Phalloidin (Invitrogen) and 20 µg/ml DAPI in 5 % FBS in PBSTx were added to all samples except the negative staining control samples and incubated at 4 C° overnight protected from light. Samples were washed five times with PBSTx with 60 min incubation time.

Embryos were mounted with low-melting point agarose on their side in optical imaging dishes. Agarose was covered with PBS. The zebrafish hearts were imaged with 3i CSU-W1 spinning disk using DAPI, mCherry and Alexa647 channels. Hamamatsu sCMOS Orca Flash4.0 camera and 40x Zeiss LD C-Apochromat water immersion objective was used. The images were analysed with Fiji software by measuring the thickness of myocardium, counting the number of nuclei and measuring the mCherry+ cell area.
